# Inquiry-Based Stress Reduction for Postpartum Anxiety and Stress During Armed Conflict: A Randomized Clinical Trial

**DOI:** 10.1101/2025.11.04.25339519

**Authors:** Shirly Mor, Yaron Sela, Shahar Lev-Ari

## Abstract

**Importance:** The postpartum period presents heightened vulnerability to stress and anxiety, particularly during wartime. While pregnancy interventions are well-studied, postpartum mental health requires targeted approaches. Inquiry-Based Stress Reduction (IBSR), combining mindfulness with cognitive reframing, has reduced depression and stress in various populations and could provide postpartum women with stress management tools during prolonged conflict.

**Objective:** To evaluate the effects of IBSR on anxiety and perceived stress in postpartum women during armed conflict.

**Design, Setting, and Participants:** This randomized clinical trial comprised an 8-week intervention and 4-week follow-up, conducted from June 9, 2024, to January 19, 2025. Participants included women aged 18-45 years up to 2 years postpartum with no psychiatric history, recruited via social media.

**Interventions:** Participants were randomly assigned (1:1) to either an 8-week IBSR group-based intervention or a waitlist control group. The IBSR program included weekly 3-hour group sessions and 1-hour guided home practice sessions.

**Main Outcomes and Measures:** Primary outcomes were anxiety (Generalized Anxiety Disorder 7-item scale [GAD-7]) and perceived stress (Perceived Stress Scale [PSS-10]). Secondary outcomes included emotional regulation, psychological well-being, quality of life, resilience, and perceived social support. Assessments were conducted at baseline (T0), postintervention (T1), and 4-week follow-up (T2).

**Results:** Among 105 randomized participants (mean [SD] age, 31.9 [6.2] years), 91 (86.7%) completed all assessments. Thirty percent of intervention participants (12 of 40) dropped below the clinical anxiety threshold (GAD-7 score <8) vs 7.7% of controls (5 of 65) (χ² = 9.01; P = .002). The intervention group showed greater reductions in anxiety (Cohen d = −0.55; 95% CI, −1.00 to −0.10; P < .05) and perceived stress (Cohen d = −1.09; 95% CI, −1.56 to −0.62; P < .01), maintained at follow-up. Cognitive reappraisal improved significantly (Cohen d = 0.99; 95% CI, 0.52-1.45; P < .01). No significant differences emerged for psychological well-being, resilience, quality of life, or social support.

**Conclusions and Relevance:** IBSR significantly reduced anxiety and stress in postpartum women during wartime, with effects maintained at 4-week follow-up. This intervention offers a scalable nonpharmacological approach for conflict-affected settings.

Trial Registration: ClinicalTrials.gov Identifier: NCT06490094

## INTRODUCTION

The postpartum period is a critical window for maternal mental health, encompassing profound physiological, emotional, and social adjustments that affect both mother and infant. While often associated with joy, this period can bring substantial psychological distress, including elevated anxiety and stress.^1–3^ Recent studies report that 12% to 39% of postpartum women experience clinically significant anxiety symptoms— considerably higher than in the general population.^4^ These emotional challenges, if unaddressed, may impair maternal functioning, infant bonding, and child development.^3,5^

The complexity of postpartum adjustment is intensified in conflict-affected regions. Armed conflicts introduce chronic stress, uncertainty, and trauma exposure, further exacerbating the psychological vulnerability of postpartum women.^6–8^ Disruption to daily routines, reduced access to health care, and the breakdown of social support systems can compound feelings of fear, isolation, and helplessness.^9^ A recent cross-sectional study conducted during a period of conflict found that postpartum women reported high levels of perceived stress and general anxiety, which were negatively associated with psychological well-being and quality of life.^8^ These findings highlight the urgent need for maternal mental health interventions in high-stress environments.

While pharmacological treatments may be effective, many women in the perinatal period prefer nonpharmacological options due to concerns about medication safety and adverse effects during breastfeeding.^10,11^ As a result, there is growing interest in psychosocial interventions that are scalable, accessible, and grounded in emotion regulation strategies and cognitive restructuring.^12,13^

Inquiry-Based Stress Reduction, developed by Byron Katie, is a structured cognitive intervention that combines mindfulness with systematic inquiry into stressful thoughts. The IBSR has demonstrated efficacy in reducing stress, depression, and anxiety and enhancing emotional regulation and well-being in various populations, including patients with cancer, caregivers, and professionals at risk of burnout.^14–24^ However, its application among postpartum women, especially in the context of prolonged stress such as war, remains underexplored.

This study aimed to evaluate the effectiveness of an 8-week IBSR intervention in reducing anxiety and perceived stress among postpartum women during an armed conflict. We hypothesized that participants in the intervention group would demonstrate significantly greater improvements than controls across these domains.

## METHODS

### Study Design and Participants

This randomized clinical trial was conducted between June 9, 2024, and January 19, 2025, and is reported in accordance with the Consolidated Standards of Reporting Trials (CONSORT) reporting guideline. The institutional review board at Tel Aviv University approved the trial (0008535-2). All participants provided written informed consent. The trial protocol and statistical analysis plan are provided in Supplement 1.

Participants were recruited via social media platforms. Inclusion criteria were mothers aged 18 to 45 years, up to 2 years postpartum, with no history of psychiatric disorders or prior exposure to the IBSR technique. Consenting participants were randomly assigned (1:1) to either the IBSR intervention or control group using the RAND function in Microsoft Excel. The allocation sequence was generated and implemented by the study coordinator (S.M.), who also enrolled participants.

The CONSORT Flow Diagram of the randomization and participant allocation process is presented in Figure 1. A total of 237 eligible participants were recruited in 2 cohorts (May and October 2024). After allocation and contact, 105 participants were retained (40 in the intervention group and 65 in the control group). Recruitment was stopped once more than 20 participants were retained in each intervention cohort, in accordance with power analysis estimates.

**Figure 1.**
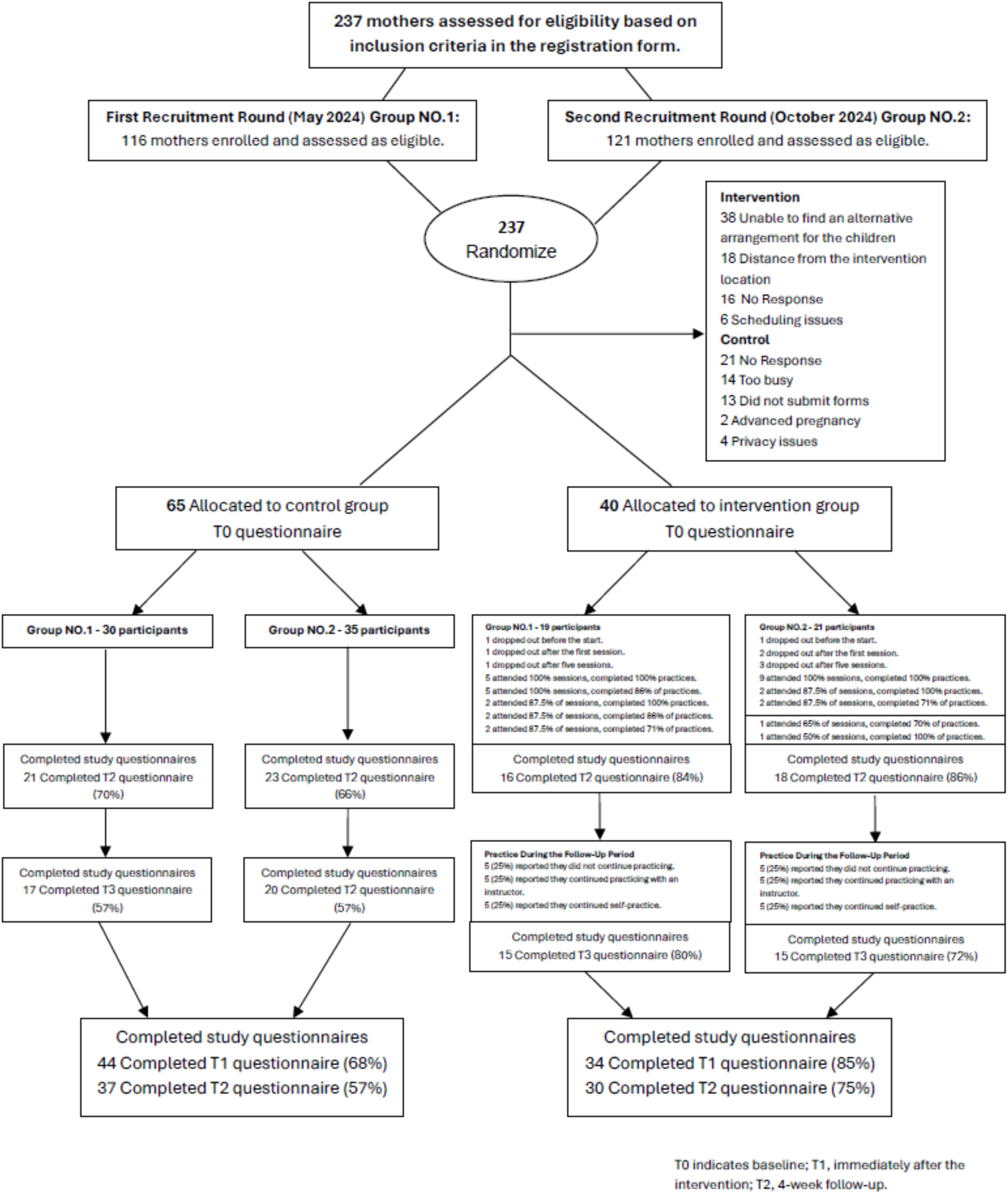
CONSORT Flow Diagram of Participant Enrollment, Randomization, Allocation, Follow-up, and Analysis. This diagram illustrates the flow of participants throughout the randomized clinical trial. A total of 237 eligible postpartum women were screened across two recruitment waves (May and October 2024). Participants were randomized in a 1:1 ratio to either the IBSR intervention group or the waitlist control group using the Excel RAND function. Following attrition, 105 participants were retained (40 in the intervention group, 65 in the control group), and 91 completed all assessments (94% in the intervention group, 88% in the control group). All randomized participants were included in the intention-to-treat analysis.

### Intervention

The 8-week IBSR program is based on The Work method, developed by Byron Katie.^24,25^ The intervention involved weekly group meetings (3 hours per meeting) throughout 8 weeks. Home practice between sessions was supported by facilitator assistants (1-hour session per week), totaling 31 hours of guided intervention. All sessions were standardized according to IBSR certification program guidelines.^26^ The intervention was conducted in 2 groups with 19 to 21 participants each. Sessions were guided by facilitators trained in the authorized certification program according to guidelines of The Institute for The Work.^27^ Timeline Design of the IBSR Intervention Program is presented in Figure 2.

**Figure 2.**
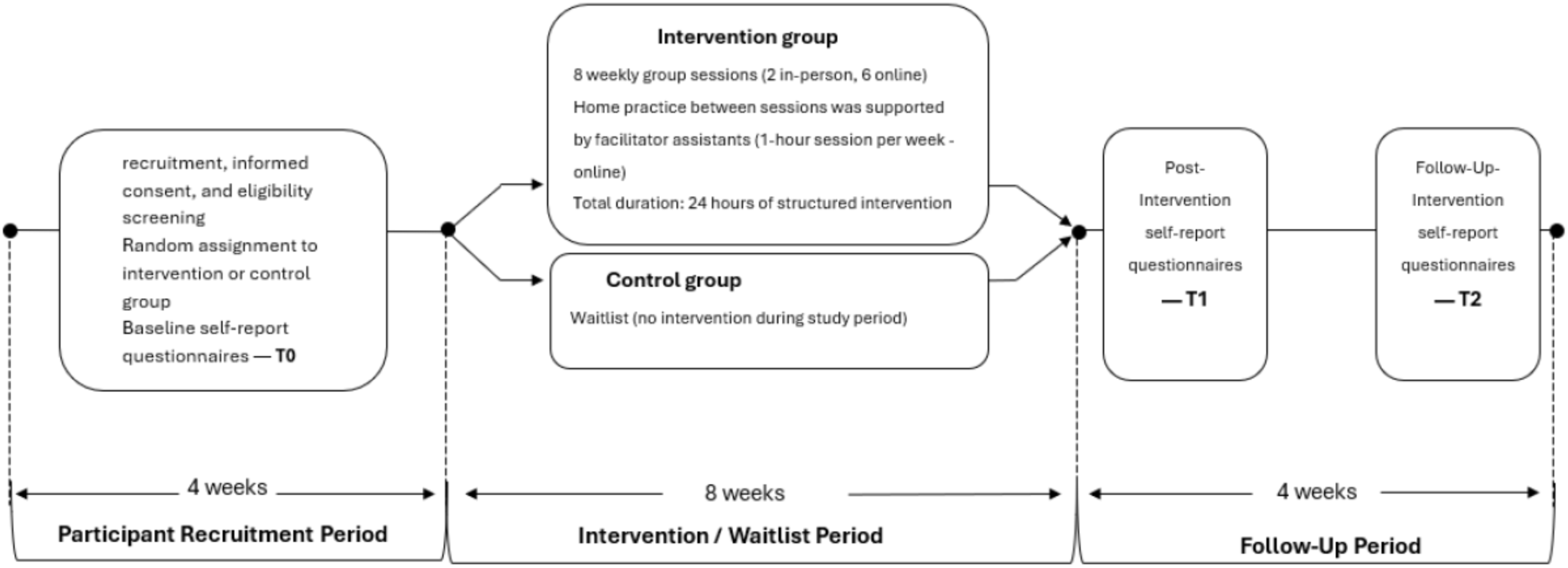
Timeline of the IBSR Intervention Program. Visual representation of the intervention design, including the 8-week group sessions, individual weekly practice, assessment points (T0, T1, T2), and allocation to intervention vs waitlist control group.

### Control

Participants in the control group were assigned to a waitlist and did not receive any intervention during the study period. They completed assessments at the same time points as the intervention group (T0, T1, and T2) and received a book on IBSR at study completion.

### Study Outcomes and Measures

Participants completed standardized self-report questionnaires at baseline (T0), postintervention (T1), and 4-week follow-up (T2).

Primary outcomes were perceived stress measured by the 10-item Perceived Stress Scale (PSS-10; scores range from 0-40, with higher scores indicating greater stress)^28^ and anxiety measured by the 7-item Generalized Anxiety Disorder scale (GAD-7; scores range from 0-21, with higher scores indicating more severe anxiety).^29^

Secondary outcomes included psychological well-being (18-item Ryff Psychological Well-Being Scale; scores range from 18-126),^30,31^ quality of life (5-item World Health Organization Well-Being Index; scores range from 0-25),^32^ emotional regulation (10-item Emotion Regulation Questionnaire; scores range from 10-70),^33^ resilience (2-item Connor-Davidson Resilience Scale; scores range from 0-8),^34^ and perceived social support (12-item Multidimensional Scale of Perceived Social Support; scores range from 12-84).^35^

Given the conflict context, exploratory measures included symptoms of posttraumatic stress disorder based on DSM-5 criteria^36^ and levels of exposure to war-related events. Participants also completed a background questionnaire covering sociodemographic variables and obstetric history. All measures demonstrated good internal consistency (Cronbach α range, 0.70-0.95; eMethods in Supplement 2).

### Statistical Analysis

Sample size calculation for the RCT was conducted using G*Power version 3.1.9 ^37^. Based on previous studies ^2,38^, a medium effect size was expected (Cohen’s d = 0.6) on the primary outcome following the IBSR intervention (T2). To detect a medium effect size with 80% power and a 5% two-sided significance level, the required sample size was 28 participants per group. To account for an expected 20% dropout rate, a total of 68 participants (34 per group) was planned.

All data were analyzed using SPSS version 28 (IBM). First, descriptive statistics were computed for all variables (means and standard deviations for continuous variables, frequencies and percentages for categorical variables). Normality of the distribution of the main psychological measures was tested using the Shapiro-Wilk test and found to be approximately normal (p > .05). Baseline group differences in sociodemographic characteristics were examined using χ^2^ tests for categorical variables (e.g., religiosity) and independent-samples t tests for continuous variables (e.g., age). Psychological outcome measures at baseline were also compared using independent-samples t tests. Variables that differed significantly between groups were included as covariates in subsequent analyses.

Prior to testing intervention effects, missing data patterns were examined using Little’s MCAR test (Little, 1988). Missing values were addressed using multiple imputation with 10 iterations. The algorithm applied multivariate regression to predict each missing value based on all other variables in the dataset, preserving the overall statistical structure of the data. The imputed values were reintegrated into the original dataset.

The analysis followed the intention-to-treat (ITT) principle, including all randomized participants regardless of dropout. To assess the intervention’s effectiveness compared to the control group, linear mixed models (LMMs) were conducted for each psychological outcome separately. The primary effect of interest was the interaction between group (intervention vs. control) and time (T0, T1, T2). Cohen’s d effect sizes with 95% confidence intervals were calculated for both within-group changes (T1 to T2) and between-group differences in change (intervention effect). All analyses were two-sided, with statistical significance set at P < .05. Data were analyzed from September 1, 2024, to February 1, 2025.

## RESULTS

### Participant Characteristics

Of 237 women contacted, 105 were randomized to the IBSR group (n = 40) or control group (n = 65) (Figure 1). The mean (SD) age was 31.9 (6.2) years; 89.5% were married, and 78.0% held vocational or academic degrees. Most deliveries were spontaneous (79.4%), and 11.4% of pregnancies were conceived via in vitro fertilization. Overall, 34.3% were primiparous.

At baseline, women in the IBSR group were older (mean [SD] age, 35.2 [4.9] vs 29.8 [6.1] years; P < .001), more often secular (26 of 40 [65.0%] vs 20 of 65 [30.8%]; P < .001), more educated (38 of 40 [95.0%] vs 34 of 65 [53.1%] with academic degrees; P < .001), had higher perceived economic status (mean [SD], 5.5 [2.2] vs 4.0 [2.5]; P = .004), shorter time since childbirth (mean [SD], 6.0 [5.6] vs 11.4 [6.4] months; P < .001), and were more often primiparous (23 of 40 [57.5%] vs 13 of 65 [20.0%]; P < .001) (Table 1). These variables were adjusted for in all analyses.

**Table 1.**
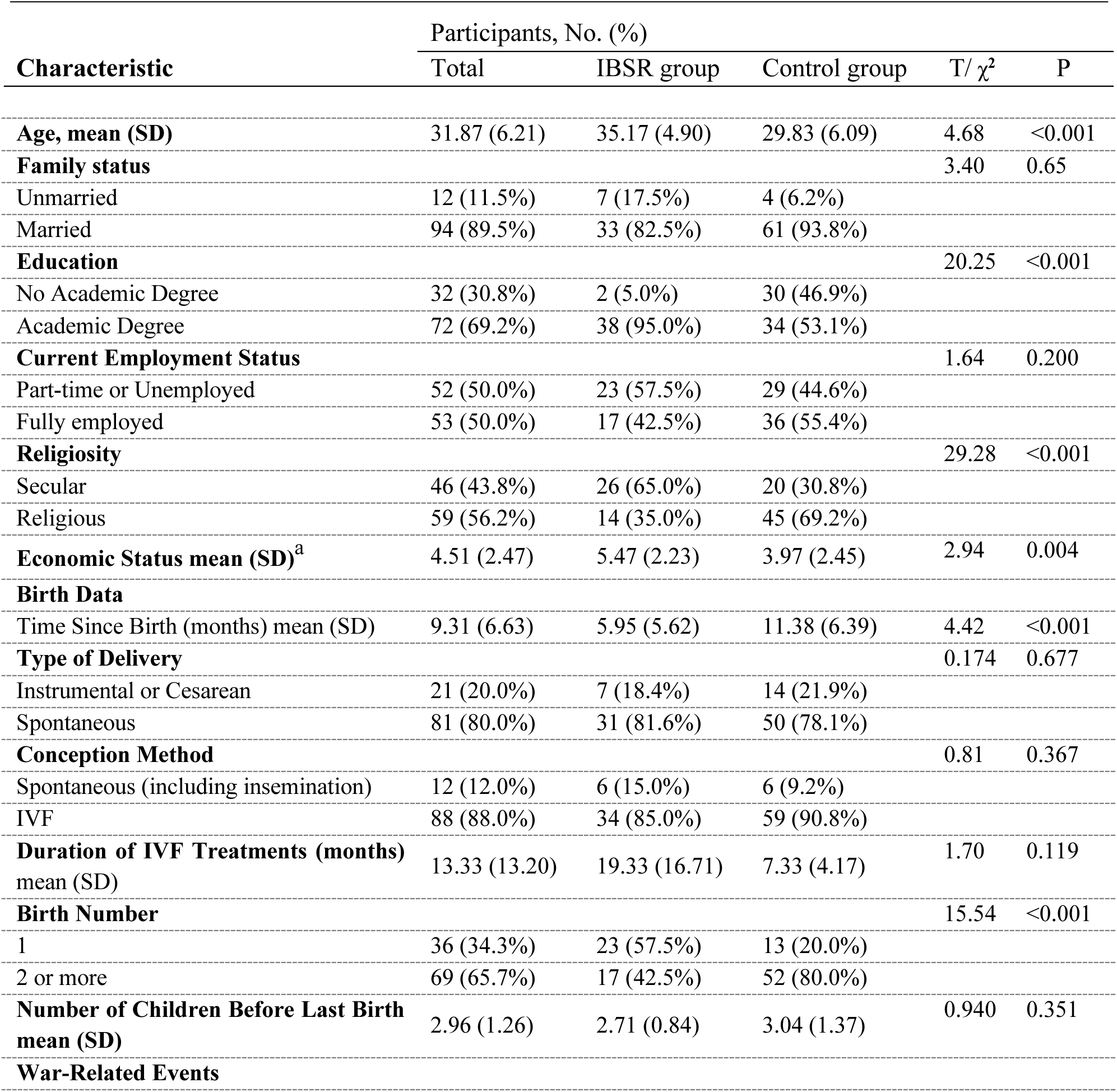
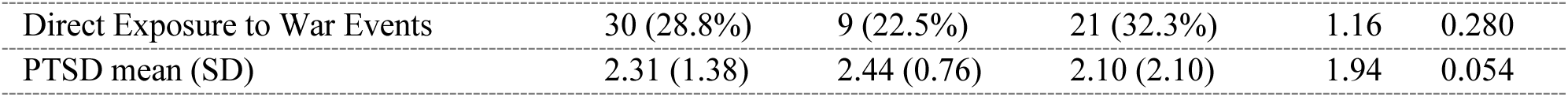
Demographic Characteristics of Participants.

| Characteristic | Participants, No. (%) | | | T/$\chi^2$ | P |
| --- | --- | --- | --- | --- | --- |
|  | Total | IBSR group | Control group |  |  |
| **Age, mean (SD)** | 31.87 (6.21) | 35.17 (4.90) | 29.83 (6.09) | 4.68 | <0.001 |
| **Family status** |  |  |  | 3.40 | 0.65 |
| Unmarried | 12 (11.5%) | 7 (17.5%) | 4 (6.2%) |  |  |
| Married | 94 (89.5%) | 33 (82.5%) | 61 (93.8%) |  |  |
| **Education** |  |  |  | 20.25 | <0.001 |
| No Academic Degree | 32 (30.8%) | 2 (5.0%) | 30 (46.9%) |  |  |
| Academic Degree | 72 (69.2%) | 38 (95.0%) | 34 (53.1%) |  |  |
| **Current Employment Status** |  |  |  | 1.64 | 0.200 |
| Part-time or Unemployed | 52 (50.0%) | 23 (57.5%) | 29 (44.6%) |  |  |
| Fully employed | 53 (50.0%) | 17 (42.5%) | 36 (55.4%) |  |  |
| **Religiosity** |  |  |  | 29.28 | <0.001 |
| Secular | 46 (43.8%) | 26 (65.0%) | 20 (30.8%) |  |  |
| Religious | 59 (56.2%) | 14 (35.0%) | 45 (69.2%) |  |  |
| **Economic Status mean (SD)<sup>a</sup>** | 4.51 (2.47) | 5.47 (2.23) | 3.97 (2.45) | 2.94 | 0.004 |
| **Birth Data** |  |  |  |  |  |
| Time Since Birth (months) mean (SD) | 9.31 (6.63) | 5.95 (5.62) | 11.38 (6.39) | 4.42 | <0.001 |
| **Type of Delivery** |  |  |  | 0.174 | 0.677 |
| Instrumental or Cesarean | 21 (20.0%) | 7 (18.4%) | 14 (21.9%) |  |  |
| Spontaneous | 81 (80.0%) | 31 (81.6%) | 50 (78.1%) |  |  |
| **Conception Method** |  |  |  | 0.81 | 0.367 |
| Spontaneous (including insemination) | 12 (12.0%) | 6 (15.0%) | 6 (9.2%) |  |  |
| IVF | 88 (88.0%) | 34 (85.0%) | 59 (90.8%) |  |  |
| **Duration of IVF Treatments (months) mean (SD)** | 13.33 (13.20) | 19.33 (16.71) | 7.33 (4.17) | 1.70 | 0.119 |
| **Birth Number** |  |  |  | 15.54 | <0.001 |
| 1 | 36 (34.3%) | 23 (57.5%) | 13 (20.0%) |  |  |
| 2 or more | 69 (65.7%) | 17 (42.5%) | 52 (80.0%) |  |  |
| **Number of Children Before Last Birth mean (SD)** | 2.96 (1.26) | 2.71 (0.84) | 3.04 (1.37) | 0.940 | 0.351 |
| **War-Related Events** |  |  |  |  |  |
| Direct Exposure to War Events | 30 (28.8%) | 9 (22.5%) | 21 (32.3%) | 1.16 | 0.280 |
| PTSD mean (SD) | 2.31 (1.38) | 2.44 (0.76) | 2.10 (2.10) | 1.94 | 0.054 |

Participants in the IBSR group reported higher perceived stress (mean [SD] PSS score, 24.1 [4.8] vs 21.3 [5.9]; P = .009) and lower quality of life (mean [SD] WHO-5 score, 9.8 [4.7] vs 11.9 [4.9]; P = .02) at baseline compared with the control group (eTable 2 in Supplement 2). No significant differences were found between participants who completed the study and those who dropped out (eTable 1 in Supplement 2).

### Primary Outcomes

A significant time × group interaction was found for anxiety (F= 3.21; P = .04), with an intervention effect size of 0.84 (95% CI, 0.20-1.46). Anxiety levels significantly decreased in the intervention group following the program (Cohen d = −0.55; 95% CI, −1.00 to −0.10), while no significant change was observed in the control group (Cohen d = −0.25; 95% CI, −0.59 to 0.09). The control group showed deterioration in anxiety during follow-up (Cohen d = 0.68; 95% CI, 0.23-1.12) (Figure 3).

**Figure 3.**
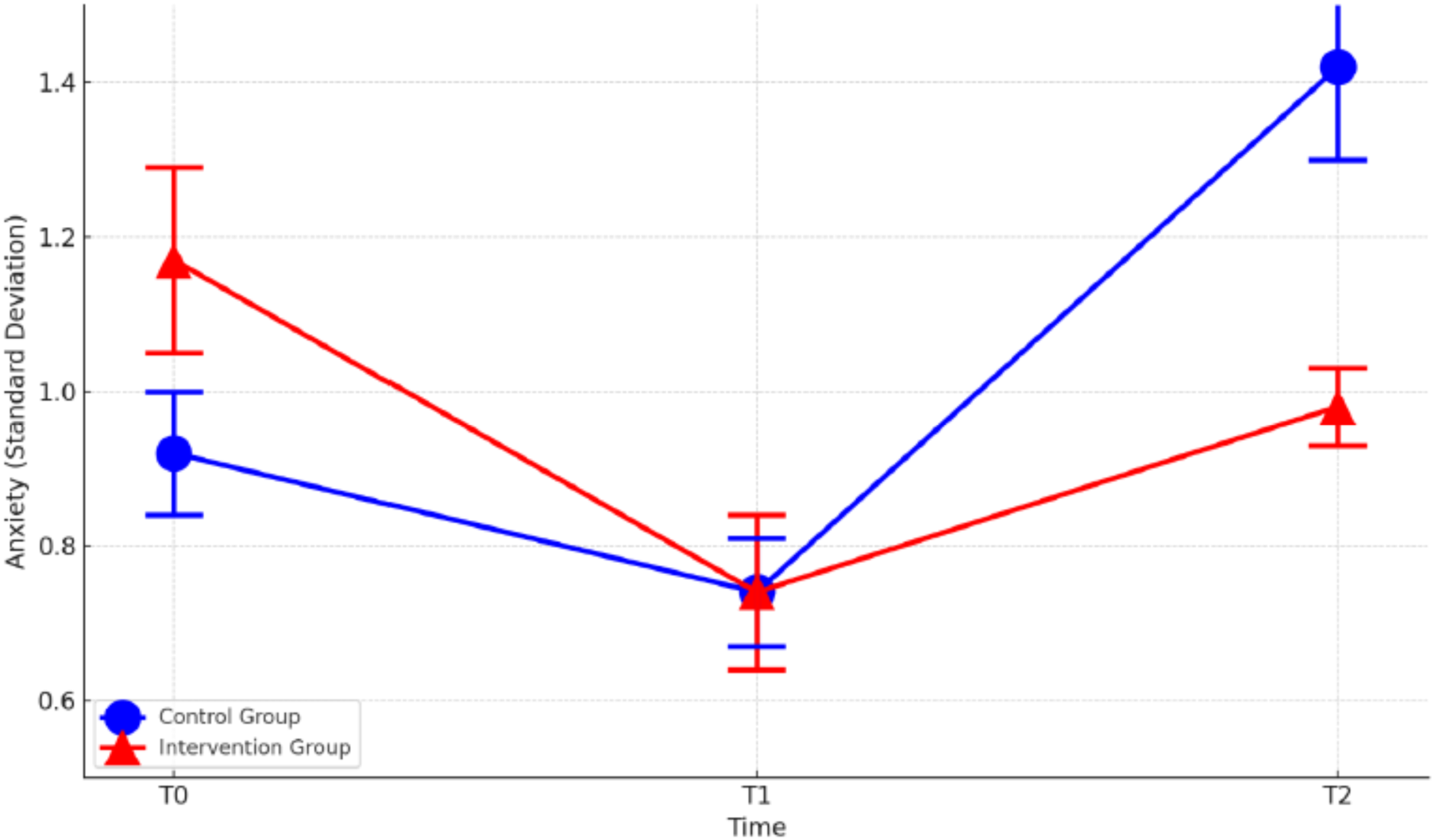
Change in Anxiety Levels Over Time by Study Group. Mean anxiety levels (GAD-7 scores) are shown at baseline (T0), post-intervention (T1), and at 4-week follow-up (T2) for the IBSR intervention group (red) and the control group (blue). Following the intervention, anxiety levels significantly decreased in the intervention group and remained stable during follow-up. In contrast, the control group showed no meaningful improvement after the intervention period and experienced a notable increase in anxiety at follow-up. Error bars represent standard deviations.

Clinical threshold analysis revealed that 30% of intervention participants (12 of 40) dropped below the clinical cutoff for anxiety (GAD-7 score <8) compared with 7.7% of controls (5 of 65) (χ^2^ = 9.01; P = .002).

A significant time × group interaction was also found for perceived stress (F₁,₆₇ = 4.95; P = .008), with an intervention effect size of 0.93 (95% CI, 0.29-1.56). The intervention group showed a large reduction in stress levels postintervention (Cohen d = −1.09; 95% CI, −1.56 to −0.62), sustained at follow-up (Cohen d = −0.85; 95% CI, −1.45 to −0.24). The control group exhibited a moderate reduction only immediately postintervention (Cohen d = −0.42; 95% CI, −0.77 to −0.07), which was not maintained (Figure 4).

**Figure 4.**
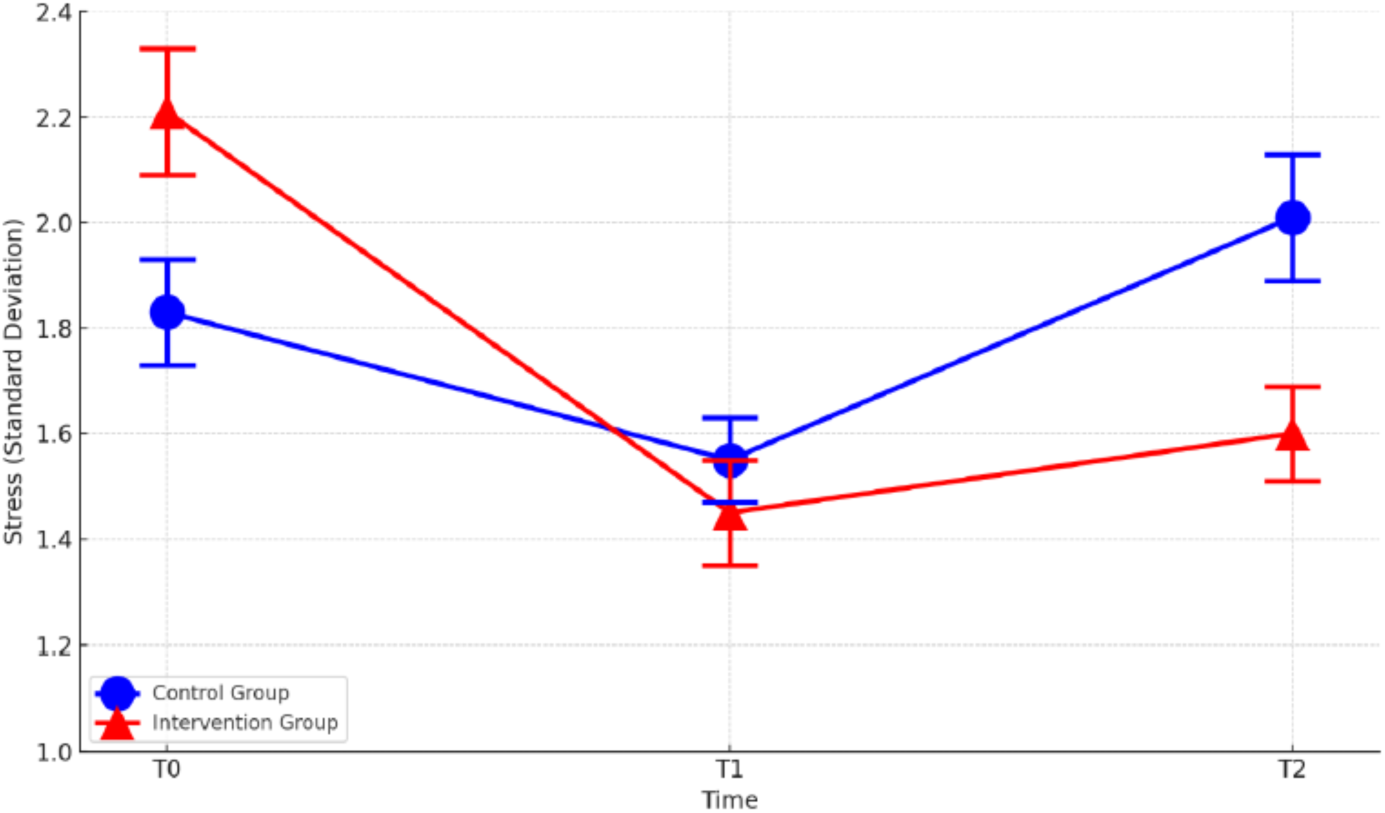
Change in Perceived Stress Over Time by Study Group. Mean levels of perceived stress (PSS) are shown at baseline (T0), post-intervention (T1), and at 4-week follow-up (T2) for the IBSR intervention group (red) and the control group (blue). The intervention group demonstrated a substantial reduction in stress immediately after the intervention (T1), with effects sustained at follow-up. The control group showed a modest temporary decrease at T1, followed by a rebound in stress levels at T2. Error bars represent standard deviations.

### Secondary Outcomes

A significant interaction was found for cognitive reappraisal (F = 4.53; P = .01), with an intervention eOect size of 0.67 (95% CI, 0.05-1.29). The intervention group showed large improvement in reappraisal postprogram (Cohen d = 0.99; 95% CI, 0.52-1.45), maintained at follow-up (Cohen d = 0.37; 95% CI, −0.21 to 0.95). No significant change occurred in the control group postintervention (Cohen d = 0.26; 95% CI, −0.08 to 0.61), with decline at follow-up (Cohen d = −0.41; 95% CI, −0.85 to −0.02).

No significant time × group interactions were found for psychological well-being (F₁,₆₇ = 0.88; P = .35), expressive suppression (F₁,₆₇ = 0.02; P = .98), resilience (F₁,₆₇ = 0.14; P = .87), quality of life (F₁,₆₇ = 1.51; P = .22), or perceived social support (F₁,₆₇ = 0.21; P = .81) (Figure 5; eTable 3 in Supplement 2).

**Figure 5.**
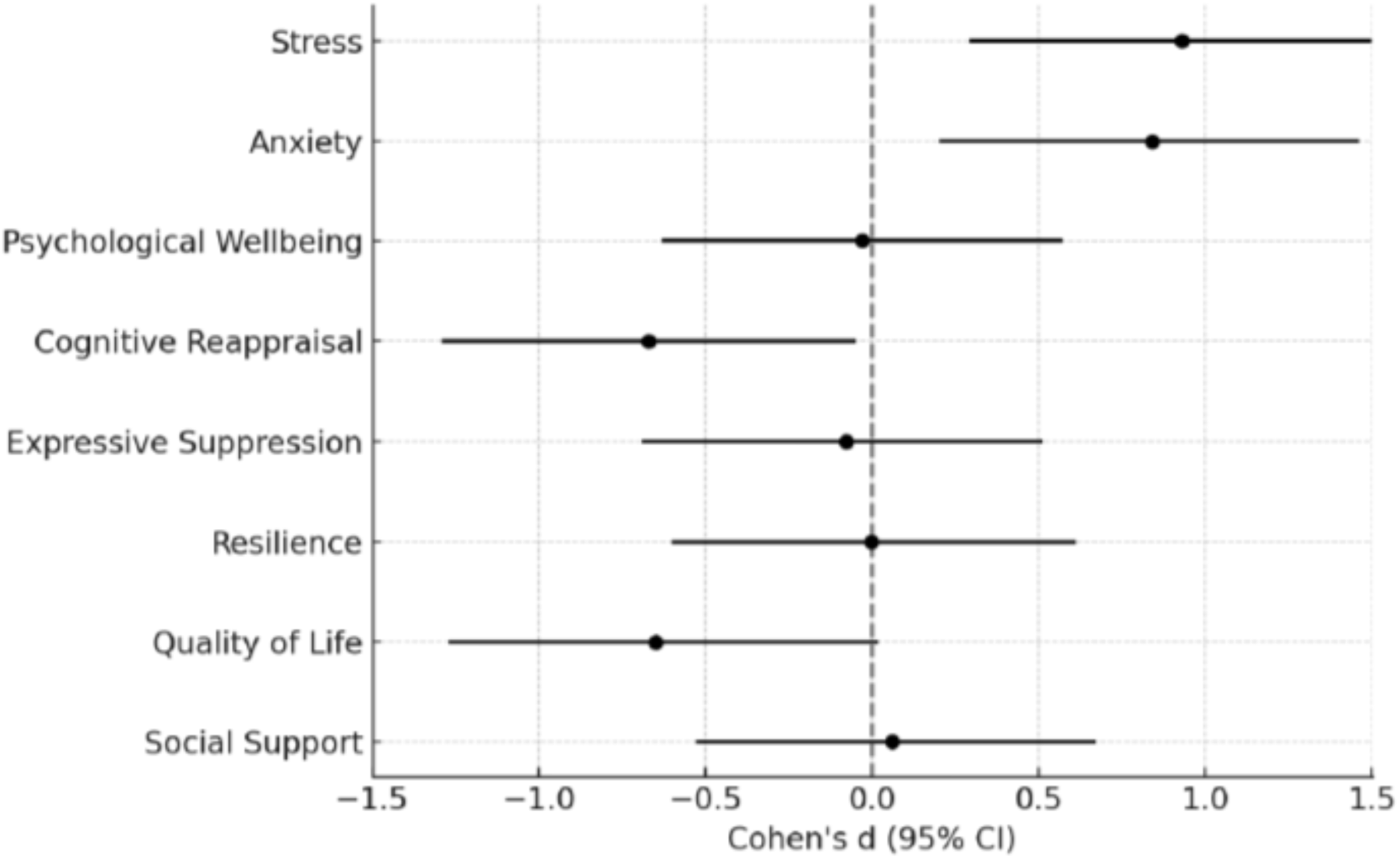
Forest Plot of Between-Group E7ect Sizes (Cohen’s d) for Primary and Secondary Outcomes. Forest plot showing standardized mean di3erences (Cohen’s d) with 95% confidence intervals for the intervention e3ect (IBSR vs control group) on each psychological outcome. Positive values indicate improvement in favor of the intervention group. Stress – Perceived Stress Scale (PSS); Anxiety –The Generalized Anxiety Disorder 7-Item (GAD-7); Psychological Wellbeing– Psychological Wellbeing (PWB); Cognitive Reappraisal/Expressive Suppression – Emotion Regulation Questionnaire (ERQ); Resilience– The Connor-Davidson Resilience Scale (10-item CD-RISC); Quality of Life – psychological general well-being by the (WHO-5), Social Support –Multidimensional Scale of Perceived Social Support (MSPSS).

## DISCUSSION

This randomized clinical trial demonstrates that an 8-week IBSR intervention significantly reduced perceived stress and anxiety among postpartum women during ongoing armed conflict. The intervention produced clinically meaningful improvements, with 30% of intervention participants falling below clinical anxiety thresholds vs 8% of controls.

The reduction in stress and anxiety was maintained 4 weeks after intervention completion, indicating short-term stability of effects despite the relatively brief program duration. Notably, the intervention group continued practicing IBSR techniques independently during the follow-up period, suggesting that participants acquired sustainable self-regulation skills. These findings support the hypothesis that self-inquiry and reframing of stressful thoughts may enhance postpartum women’s coping capacity during periods of heightened psychological vulnerability.^9^

The improvement in emotional regulation through cognitive reappraisal suggests that IBSR may not only alleviate distress symptoms but also promote adaptive emotional regulation strategies—consistent with previous research.^20,24^ Such strategies are critical for healthy adjustment to dynamic challenges in the postpartum period, marked by significant changes in personal, familial, and social functioning. The improvement in cognitive reappraisal aligns with theoretical models suggesting that IBSR fosters emotional adaptation through structured self-inquiry and cognitive reframing.^14–24^ This particularly benefits postpartum women who report feelings of guilt, doubt, and self-criticism. The nonpathological and accessible nature of IBSR—requiring no diagnosis, medication, or therapeutic experience—may especially empower women during this demanding period.

Despite significant effects on stress, anxiety, and cognitive reappraisal, no significant changes occurred in psychological well-being, resilience, quality of life, or perceived social support. This raises questions about these protective factors’ short-term sensitivity. Often conceptualized as stable traits shaped gradually through internal and environmental resources,^40–44^ previous IBSR studies reported improvements even after brief interventions.^16,18,22^ Cultural context, participant characteristics, or intervention focus—emphasizing stress and anxiety reduction over well-being and resilience enhancement—may explain divergent outcomes. Postpartum mothers navigating intense transitions may require immediate distress relief before longer-term well-being, resilience, or support shifts emerge.^1,44^ Stress and emotional regulation improvements may represent critical early steps, potentially enabling broader psychological growth.

The unique context of this study—during active armed conflict—plays a central role in understanding the findings. National emergencies, especially prolonged conflicts, are major risk factors for maternal mental health, heightening anxiety, stress, and posttraumatic stress disorder symptoms during the perinatal period.^46–48^ Studies from Ukraine during wartime highlight the unique vulnerability of postpartum women in prolonged emergencies, showing that even when acute distress symptoms improve, broader indicators such as well-being and quality of life often remain unchanged.^6,49^ Similarly, research during the COVID-19 pandemic found that while anxiety and stress increased, measures of psychological well-being and resilience remained relatively stable.^50,51^ These patterns suggest that in conflict-affected populations, improvements in broader psychological outcomes may require longer-term stabilization processes beyond immediate symptom-focused interventions.

### Limitations

This study has several limitations. First, despite randomization, significant baseline sociodemographic differences emerged between groups. Although statistically controlled, these may reflect access or motivation differences suggesting selection bias. Second, self-reported data introduces potential social desirability or recall bias. Third, the 4-week follow-up limits conclusions about long-term effects. Finally, the sample’s high education level and above-average income reduce generalizability.

## Conclusions

This randomized clinical trial found that IBSR was effective for reducing anxiety and stress and enhancing emotional regulation in postpartum women in an armed conflict region. Given the rising prevalence of postpartum psychological distress, particularly in conflict-affected regions, there is an urgent need for accessible, nonpharmacological interventions that can support maternal mental health. The IBSR may serve as a meaningful tool within postpartum mental health care, offering a structured approach to alleviating acute distress and strengthening emotional coping capacities.

## Data Availability

Deidentified individual participant data and statistical code are available on reasonable request from the corresponding author.

## Additional Contributions

We thank Michal Evrach-Bar, PhD (Chair), and Bruria Adini, PhD, of the supervisory committee for their guidance and support. We thank the certified IBSR facilitators who volunteered their time and expertise. We are grateful to all participating mothers for their engagement and trust. No compensation was provided for these contributions.

